# Study protocol: Lead exposure, diet, and cognitive development among school-age children in Guayaquil, Ecuador

**DOI:** 10.64898/2026.01.28.26345079

**Authors:** Marcelo Armijos Briones, Elisa Iturralde Brinkmann, Dana Milena Crespo Párraga, Patricia Andrea Uscocovich Jiménez, Diana Lucía Tinoco Caicedo, Paola Calle-Delgado, Christian Moreno Alvarado, Nicole Armijos Bazurto, Diana Glicelia Tomalá Castro, Paula Viktoria Báez Freire, Emily Angeline Zambrano Mendoza, Doménica Barcia Roca, Brenda Angélica Bucheli Bermúdez

**Affiliations:** School of Dentistry. Universidad de Especialidades Espíritu Santo, Samborondón, Ecuador; Facultad de Ciencias Naturales y Matematicas (FCNM). Escuela Superior Politécnica del Litoral, Guayaquil, Ecuador; PVALOR S.A.S.; Faculty of Humanities. Universidad de Especialidades Espíritu Santo, Samborondón, Ecuador

## Abstract

Lead exposure is a major environmental health concern, especially in childhood, due to its neurotoxic effects even at low levels. This cross-sectional observational study aims to evaluate the association between lead accumulation in tooth enamel, cognitive performance, and dietary patterns in children aged 7 to 9 years belonging to the Archdiocesan Educational Network of Guayaquil, Ecuador. A total of 384 participants will be selected using stratified sampling according to socioeconomic status. Lead levels will be assessed using a validated dental enamel biopsy technique, while IQ will be measured using the Wechsler Intelligence Scale for Children – Fifth Edition (WISC-V). Dietary intake will be assessed using a Food Frequency Questionnaire (FFQ) validated for the Ecuadorian population. All procedures will ensure the safety, comfort, and confidentiality of participants. Potential risks, such as emotional distress or academic disruption, will be minimized through adapted protocols and ethical safeguards. This study seeks to provide critical local evidence on environmental lead exposure and its association with cognitive outcomes and dietary, to help inform future public health efforts.

## Introduction

Lead exposure is a serious environmental health problem, especially in childhood, due to its harmful effects on cognitive and neurological development. This toxic metal accumulates mainly in hard tissues of the body, such as bones and teeth, where it can remain for decades, has been associated with cognitive impairment and lower IQ scores, even at exposure levels considered low [1–3]. Several studies have shown that there is no safe level of lead in the human body and that minimal exposure can affect brain development from early stages of life, interfering with essential neural functions and increasing the risk of behavioral disorders such as aggression or antisocial behavior [4–6].

Lead contamination can come from different sources, the main ones being environmentally and dietary [7]. Exposure to this metal and its levels can be detected with different biological markers such as blood, bones, teeth, saliva, hair, and nails [8]. Blood is the most effective marker for correlating lead levels in recent exposures. To determine the presence of lead in past exposures, hard tissues such as bone and teeth are more effective biomarkers. The use of bone presents several limitations, such as the cost and pain of biopsies, and the difficulty in finding suitable bone for determining lead levels [9]. The best option for determining lead levels due to past exposures, even in human and primate fossils, is tooth enamel [10]. Even lead that may be retained in the bone matrix can be released into the body, causing some cognitive impairments, and then be absorbed into developing teeth as tooth buds [11], giving the enamel of permanent teeth significant attributes for understanding past lead exposure.

It is important to know the location of lead in the body in relation to the time of exposure. Above all, because environmental pollution, diet, drinking water, and even the materials used to build houses can significantly increase lead levels in the body. Furthermore, it is known that this metal can be incorporated into the body even during pregnancy [12]. Measuring lead levels in children is more effective because they have stable living conditions and controlled diets, making it easier to determine whether their environment or diet is associated with lead exposure. Studies have shown that people living in densely populated urban areas tend to have higher consumption of processed and ultra-processed foods. Diets high in ultra-processed foods may also be associated with a greater risk of exposure to foodborne contaminants, including heavy metals [13,14]. For example, in Ecuador, the National Agency for Health Regulation, Control, and Surveillance (ARCSA) found levels of heavy metals exceeding Ecuadorian limits during routine quality analyses of certain food products. This was the case with powdered cinnamon ready for use in food, where three samples of ground cinnamon had a lead content higher than the maximum level established by Ecuadorian and European Union regulations (2.0 mg/kg) [15]. This set off alarms, and since then, several studies have been conducted in this country to verify the presence or absence of these types of compounds.

This is the case of studies such as the one carried out in Ecuador, which identified worrying levels of lead (Pb) in bananas grown in the provinces of Bolívar and Santa Elena. The estimated concentrations reached approximately 3.858 and 4.932 µg Pb/kg·day, respectively, exceeding the FDA’s recommended daily intake of 3 µg of lead for children [16]. Prolonged lead intake above these limits can have a significant adverse association on children’s health. It is associated with delays in cognitive development, decreased IQ, attention difficulties, and other neurobehavioral effects over time [17].

In the city of Quito, research was conducted to determine the presence of heavy metals in a popular market in the city. The research showed that several unprocessed products had high concentrations of metals, including lead and cadmium [18]. In the same city, some water sources were found to be contaminated with these same metals [18]. For its part, Ecuador’s Ministry of Public Health conducted blood tests to quantify heavy metal levels in 300 people, finding that some of them had high levels of these metals. This latest study is one of the few in which people have participated. However, it was not possible to determine the source of exposure. An international comparative study showed that the observed lead concentration levels in Ecuadorian infants and young children were 29.4 μg/dL [19], well above the current Centers for Disease Control and Prevention (CDC) reference value of 5 μg/dL [20]. This implies that they have a higher potential for metal-mediated poisoning than children of similar ages in Asia, Europe, other Latin American countries, and the United States.

In highly urbanized cities such as Guayaquil, where high levels of pollution and unfavorable socioeconomic conditions converge, children are particularly exposed to sources of lead present in the environment, soil, everyday products, and even school supplies [15,16]. Given this problem, there is an urgent need to characterize exposure levels and examine associations between lead exposure and children’s health outcomes, especially considering the vulnerability of populations in low-income areas.

The Archdiocese of Guayaquil (Catholic Church) maintained schools throughout the city of Guayaquil and grouped them into the Archdiocesan Educational Network. A total of 21 schools comprise this network, which aims to safeguard the education of children from different socioeconomic backgrounds, but primarily those from disadvantaged backgrounds. Parents contribute a small amount for the school’s operation (approximately $40 to $60 per month), receive some government incentives, and the Catholic Church covers the remaining expenses [25].

This study aims to describe the association between lead levels in tooth enamel and children’s IQ and dietary patterns in Guayaquil city. Our hypothesis is that higher lead levels will be associated with lower IQ. Furthermore, we hypothesize that there is a dietary pattern related to the amount of lead present in the tooth enamel of these children. Given the cross-sectional design, analyses will estimate associations and will not support causal inference

## Materials and Methods

A cross-sectional observational study will be conducted to examine the relationship between lead levels in dental enamel, intelligence quotient (IQ), and dietary patterns in children aged 7 to 12 years from schools within the Archdiocese Educational Network (REA) of Guayaquil. The study will include students from Josemaría Escrivá, San Francisco Javier, San Joaquín y Santana, Dolores Sopeña, and Néstor Astudillo schools, located across the north, northeast, central, and southern areas of the city. Although the sample is not fully representative of Guayaquil’s population of over three million, it includes participants from key urban zones and will provide valuable baseline data for future research in Ecuador.

The sample size was calculated considering a population defined by the number of students in the required age range at each educational unit. Stratified sampling was performed as follows:

The sample size was calculated based on the total number of students duly enrolled in the selected educational units corresponding to the selected age range, as shown in Table 1. A 95% confidence level, a 5% margin of error, and an expected frequency of 80% were used. The expected frequency was set at 80% as a conservative value, considering previous evidence on lead exposure in dental enamel in pediatric populations from similar contexts. In Brazil, studies using in vivo microbiopsy reported detectable lead concentrations in all samples analyzed, both in deciduous and permanent teeth, with ranges between 5 and 4711 μg/g [21,22]. A study in Peru found that 100% of the children evaluated had detectable levels of lead in their dental enamel (5.65–12.83 μg/g) [23]. According to data from each of the educational units, the study population consisted of 906 children. With this population, the minimum required sample size was 193 children. Dividing the calculated required sample size by the total population of children in the age range across all schools yielded a fraction of 0.21. This fraction was then multiplied by the number of children in each school to obtain the minimum sample size for each educational unit, as shown in Table 1.

**Table 1.** Distribution of the study population and proportional allocation of the minimum required sample size across participating schools.

| School | Number of children aged 6 to 12 years | Required sample size |
| --- | --- | --- |
| Josemaría Escrivá | 248 | 53 |
| Francisco Javier | 83 | 18 |
| San Joaquin y Santana | 333 | 70 |
| Dolores Sopena | 92 | 20 |
| Nestor Astudillo | 150 | 32 |
| Total | 906 | 193 |

### Eligibility Criteria

#### Inclusion Criteria

Children aged 6 to 12 years with fully erupted permanent central incisors are expected to participate in this research. Their legal representatives must have signed the informed consent form, and the children must have signed the informed assent form appropriate for their age.

#### Exclusion Criteria

Children diagnosed with molar-incisor hypomineralization or any type of enamel alteration, and children with any type of cognitive disability will not be eligible to participate. Additionally, children with delayed incisor eruptions are not eligible.

### Procedure

Data collection is planned to be carried out as follows: once the informed consent form has been completed, the link to the Google form with questions about food consumption patterns (Supplementary File 1) will be sent to the legal representatives of the children who agree to participate. Then, psychologists will visit the educational units to assess IQ, and finally, a dentist will visit the schools to perform enamel biopsies. Data collection has not yet begun. It is expected to start in January 2026 and conclude in March of the same year.

#### Collection of tooth enamel samples

The entire procedure will be performed at the facilities of the schools participating in this study. A Trophy brand portable dental chair will be set up for this purpose. First, the tooth selected for the biopsy (tooth 2.1), which will always be present, will be cleaned. Cleaning will be performed with a low-speed rotary brush. After that, the tooth will be dried for 5 seconds using the portable chair’s triple syringe. Relative isolation will be achieved using cotton rolls on the cheeks. With the tooth clean and dry, a square of Scotch 311 brown adhesive tape will be placed as shown in Figure 1a. This square of tape will have a 1.5 mm diameter perforation in the center. This differs from the reference procedure, where Scotch 810 tape, which is transparent and has a 1.6 mm diameter perforation, was used [21]. It is important to mention that, in the pilot test we conducted, it proved vital that the tape not be placed over the incisal or gingival margin, as this causes it to detach, especially during rinsing with ultrapure water. The droplet disperses and cannot be collected with the pipette tip, as shown in Figure 1b. Furthermore, when introducing the pipette tip into the container with hydrochloric acid and glycerol, the more viscous solution retains droplets on the outside of the tip, which could increase the final amount of solution. It is important to remove this excess with a clean cotton swab, as shown in Figure 1b.

**Figure 1.**
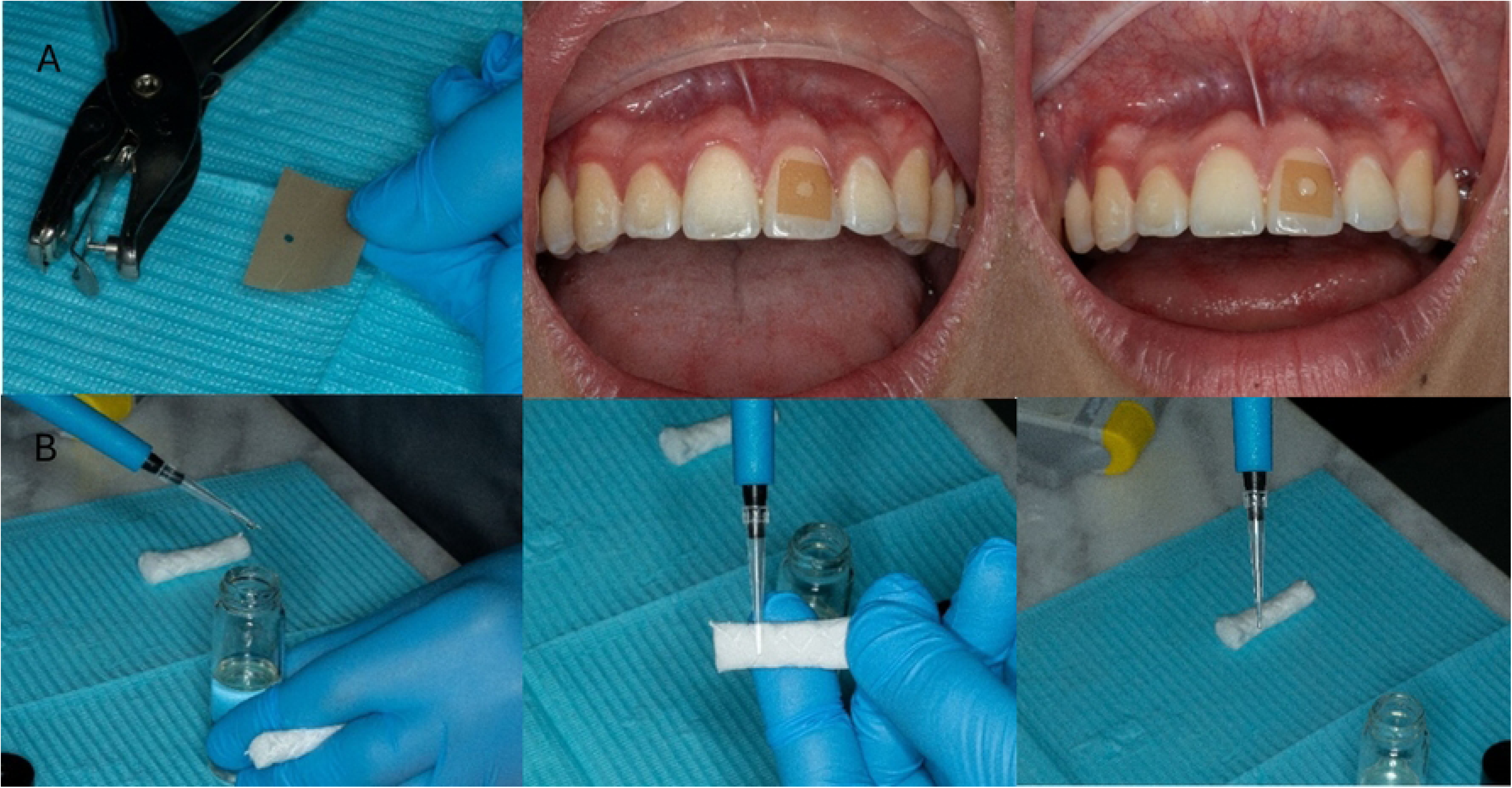
Description of the procedure prior to enamel biopsy collection. The images were taken at the clinics of Universidad Espíritu Santo and are for illustrative purposes only; they do not correspond to biopsies taken at the school with the participating children.

With the tooth prepared, 5 µL of 1.6% HCl in 70% glycerol will be applied for 20 seconds. The drop will be gently swirled with the tip of the pipette. The biopsy solution will be transferred to a 0.2 ml centrifuge tube (Axygen Scientific, Inc., Union City, USA) containing 200 µL of ultrapure water. The enamel surface will be washed with 5 µL of ultrapure water, and this wash will also be collected.

During pilot tests with extracted teeth and in 4 different individuals (in vivo), the amount of phosphorus was below the lower limit required to reach the appropriate depth for accurate measurement of lead content in the enamel. Therefore, the procedure described above is performed twice at the same site where the perforated tape will be placed. This will result in a total of 220 µL in the capped Eppendorf tube where the sample will be stored. At the end of the procedure, the tape will be removed, the tooth will be dried, and 2% sodium fluoride (Eufar brand) will be applied. The coded sample will be stored in a thermal container and then transported the same day to the laboratory where the samples will be processed.

#### Chemical Analysis

The chemical analysis for the determination of lead in the enamel samples will be carried out following the procedure described in a previous study [21], with specific adaptations to the available equipment. After collecting the enamel biopsies, the samples will be dehydrated under vacuum with anhydrous calcium chloride for 36 hours to prevent volume loss due to evaporation. Subsequently, they will be rehydrated with 220 µL of ultrapure water and divided into two aliquots: one for lead determination and the other for phosphorus.

The lead concentration will be determined by graphite furnace atomic absorption spectrometry using a Perkin Elmer Thermo Scientific Genesys 20 UV-vis atomic absorption spectrophotometer, manufactured in 2006. The samples will be diluted to a final volume of 490 µL with a solution containing 0.2% (w/v) NH₄H₂PO₄, 0.5% (v/v) Triton X-100, and 0.2% HNO₃, following the recommendations of the original protocol.

All materials used will be pre-washed with 10% nitric acid overnight and rinsed with ultrapure water before use to prevent contamination by traces of lead. A certified reference material (SRM 1640, from the National Institute of Standards and Technology) will be used to verify the accuracy of the analytical method.

#### Saliva sample (for future research on bacteria resistant to lead)

The objective of this part of the procedure is to collect a saliva sample from the children participating in the research to detect the presence of bacteria in the oral cavity of those with elevated lead levels and to try to identify any genetic variations in the bacteria that help them resist the presence of the metal.

To do this, the children will be asked to provide an unstimulated saliva sample in a sterile 2 ml Eppendorf tube with a cap, which will be transported to the facilities of the Universidad de Especialidades Espíritu Santo and stored in a freezer capable of reaching temperatures as low as -80 degrees Celsius.

### Intelligence quotient assessment

#### Intelligence quotient assessment

The Wechsler Intelligence Scale for Children - Fifth Edition (WISC-V) will be used. This is an internationally standardized clinical instrument, designed for individual use, which assesses the intellectual abilities of children between 6 years and 0 months and 16 years and 11 months. The WISC-V scale consists of fifteen subtests. For this assessment, an abbreviated version will be administered that includes the ten main subtests: Blocks, Similarities, Matrices, Digits, Cues, Vocabulary, Scales, Visual Spatial, Picture Completion, and Symbol Search. This selection provides a representative estimate of intellectual ability and a comprehensive assessment of the five main indices of the WISC-V scale: Verbal Comprehension Index, Visual-Spatial Index, Fluid Reasoning Index, Working Memory Index, and Processing Speed Index. Each of these indices assesses a specific cognitive ability through different subtests and, together, they provide the total IQ score. The full scale IQ (FSI) is a composite score that represents an overall estimate of cognitive functioning. In the WISC-V, it is calculated from the five main indices and seeks to reflect an individual’s overall ability to reason, solve problems, understand complex information, and adapt to the environment. This score is interpreted within a standardized scale in which scores between 90 and 109 are considered average, with a minimum score of 45 and a maximum of 155.

The tests will be administered by four psychologists duly authorized to practice in Ecuador. These evaluations will be administered in the library or in an available break room within the educational units, in coordination with the authorities of each school. A maximum of 25 children will be evaluated per day. To avoid fatigue or tiredness among participants, the test will be divided into three one-hour sessions on consecutive days.

On the first day, the evaluation group will be introduced to the participants, with the aim of building trust among the children to facilitate the evaluation process. Next, the subtests corresponding to the verbal and visuospatial comprehension scales will be administered. The first subtest to be administered will be the Cubes subtest, which corresponds to the visuospatial scale, in which the child is presented with an image that they must reproduce with two-colored cubes within a certain time. Next, the Similarities subtest of the Verbal Comprehension scale, which consists of the evaluator reading two words that represent objects or concepts familiar to the child, who must describe their similarity. Third, the Vocabulary test from the Verbal Comprehension scale consists of graphic questions in which the child must respond with the name of the images shown and verbal questions in which they must define the words read by the evaluator. Finally, the Visual Puzzles from the Visual-Spatial scale will be administered. In this subtest, the evaluator presents the child with a completed puzzle, and the child must select three answer options that allow them to reconstruct it within a certain time.

On the second day, the fluid reasoning and working memory scales will be administered. We will begin with the fluid reasoning matrices subtest, in which the child is presented with an incomplete matrix and must select the answer option that completes it correctly. Next, working memory is assessed using the digit subtest, in which the evaluator reads a series of numbers to the child, who must repeat them in order. This test consists of three tasks: (1) Digits in direct order, in which the child repeats the numbers in the same order in which they are heard; (2) Digits in reverse order, in which the child repeats the numbers in reverse order to the order in which they are heard; and (3) Digits in ascending order, in which the child repeats the numbers in ascending order. Next, for fluid reasoning, the balance test will be administered, which consists of presenting the child with an image of a balance with one or more weights missing and asking them to select the answer option that keeps the balance balanced within a certain time. To conclude the second session, the working memory scale picture scanning test will be administered. For this test, the child will look at a page of stimuli and then select those they have already seen from the options presented by the evaluator on the second page.

During the third and final day of assessment, the processing speed scale will be administered using the signals and symbol search subtests. The signals require the child to use a key within a certain time to copy the correct symbols. The signals require the child to press a key within a certain time to copy the symbols corresponding to the numbers or figures presented. Symbol searching, on the other hand, requires the child to indicate whether any of the target symbols are present in the search set. Finally, subjects will be given positive emotional feedback to conclude the assessment process and thank them for their participation.

To ensure the child’s safety, privacy, and comfort, they will always be accompanied by the evaluator and a representative from the school’s psychology department. In the event of fatigue or signs of distraction on the part of the child, a short break will be allowed between subtests. The information obtained will be stored in encrypted format, respecting the confidentiality of the participants evaluated.

### Assessment of dietary patterns

One of the objectives of this research is to associate food consumption patterns with the possible presence and quantity of lead in the dental enamel of participating children. To this end, the Food Quality Questionnaire, created by the Global Food Quality Project, will be used. This questionnaire consists of 29 yes/no questions about the meals consumed the day before completion. The questions have been developed and adapted to the Ecuadorian population, including foods and beverages characteristic of the diet [24]. To facilitate these questions, boxes were included in the questionnaire for respondents to mark “yes”; if no box is marked, it indicates that the food was not consumed. In addition, questions were developed to gather information on the consumption of foods and the use of kitchen utensils that, according to scientific evidence and the knowledge of some consulted experts, may contain lead, hinder or facilitate its absorption. Both questionnaires were combined into one, as shown in Supplementary File 1. Questions 1 through 6 address social variables. Questions 7 through 14 correspond to the Food Quality Questionnaire, and questions 15 through 37 to the new section created for this study. In total, there are 37 questions with an average completion time of 15 minutes.

#### Questionnaire on the frequency of consumption of foods with potential content and ease and/or difficulty of lead absorption

This section is designed to assess common dietary patterns and contextual factors that may influence lead exposure or modulate lead absorption in children. Each subsection covers a distinct but interrelated dimension of diet-related risk or protection, based on biochemical, environmental, and epidemiological evidence.

#### Potential Pathways of Dietary Lead Exposure

This section includes food groups most likely to contribute to dietary lead intake through contamination of raw materials, soil, water, or food processing. Several studies have identified cereals, vegetables, tubers, and pulses as major sources of dietary lead exposure in children. In a recent exposure assessment, cereals and baked goods were the leading sources of lead intake in infants and young children in the United States [25]. Similar results were observed in South American populations, where dietary lead exposure in children was dominated by cereals, vegetables, and tubers grown in contaminated soils [26]. Root and leafy vegetables efficiently accumulate lead through irrigation water and soil, while fish, shellfish, and animal organs can bioaccumulate it via aquatic or trophic pathways [27]. In addition, informal and artisanal foods, such as street food, sweets, and homemade condiments, represent context-specific sources of lead contamination due to the use of low-quality colorings, unregulated ingredients, or contact with lead-glazed utensils[28]. Processed and canned foods can contribute to lead contamination through soldered packaging or migration from metal surfaces, and ice or beverages made with untreated water constitute indirect exposure pathways in areas with poor drinking water.

#### Dietary Modulators of Lead Absorption (Protective and Susceptibility Factors)

Previous research has determined that adequate intake of calcium, iron, and vitamin C reduces gastrointestinal lead absorption by competing for transporters and enhancing antioxidant defense. Increased calcium intake significantly lowers blood lead levels in exposed children [29], while iron deficiency increases absorption via Divalent Metal Transporter 1 (DMT-1) mediated pathways [30]. Vitamin C enhances the absorption of non-heme iron and directly chelates lead ions, further reducing systemic absorption [31].

Conversely, tea and coffee consumed with meals decrease the bioavailability of iron and calcium due to tannins and polyphenols, which may increase susceptibility. Regular fasting or skipping meals has been associated with higher blood lead concentrations, as the solubility and intestinal absorption of lead increase on an empty stomach [32].

#### Water and Cooking Utensils (Contextual Environmental Factors)

While lead exposure is primarily environmental, dietary pathways interact with household and food preparation practices. Drinking and cooking water can be a significant source of lead when distributed through older pipes, unlined tanks, or contaminated wells [33,34] Measuring the domestic water source (public network, well, cistern, bottled, boiled, or filtered) allows for differentiation between potentially contaminated and safer supplies. Lead-glazed ceramics and aluminum cookware have also been identified as preventable sources of chronic dietary lead exposure in Latin America and other low- and middle-income countries [35]. Acidic foods prepared or stored in such containers promote lead leaching. Therefore, questions about the use and frequency of ceramic or aluminum cookware were included to encompass these exposure contexts.

#### Eating Places and Food Environment

This section addresses the environmental dimension of dietary exposure by identifying where and how food is obtained and consumed. Street food and vendors have been identified as recurring sources of contamination due to environmental deposition of lead dust, the use of unsafe cookware, and unregulated handling of ingredients [28].

##### Content Validity

The relevance and clarity of the items in the Question Block on the consumption of foods with potential lead content and those with a potential protective association against lead absorption were evaluated by four experts from different specialties: a nutritionist, a gastroenterologist and nutritionist, an epidemiologist, and a physician with knowledge of toxicology. Each item was rated on an ordinal scale of 1 to 4 for Relevance (1 = not relevant, 4 = very relevant) and Clarity (1 = confusing, 4 = very clear). The Content Validity Index (CVI) for each item was calculated as the proportion of experts who rated each question with scores of 3 or 4 for relevance, and the average Content Validity Index (S-CVI/Ave) was calculated as the means of all the CVI scores for relevance. The I-CVI values for relevance ranged from 0.75 to 1.00, while those for clarity ranged from 0.50 to 1.00. The overall S-CVI/Ave was 0.85, indicating acceptable content validity according to the criteria of Polit & Beck (2006) [36]. Table 2 shows that most items were considered highly relevant (I-CVI ≥ 0.78), especially those related to foods of animal origin, street food, and ceramic or aluminum utensils.

**Table 2.** Content validity indices (I-CVI) for each item of Block B: dietary pattern and potential lead exposure.

| No . | Item | I-CVI (Relevance) | I-CVI (Clarity) | Interpretation |
| --- | --- | --- | --- | --- |
| 1 | Leafy vegetables (lettuce, spinach, parsley) | 0.50 | 0.25 | Reformulate |
| 2 | Root or tuber vegetables (potato, cassava, carrot, beet) | 0.75 | 0.25 | Revise |
| 3 | Grains and cereals (rice, maize, bread/wheat, oats) | 0.75 | 0.25 | Revise |
| 4 | Legumes (beans, lentils, chickpeas, peas) | 0.50 | 0.25 | Reformulate |
| 5 | Fish, tuna, sardines, or seafood | 0.75 | 0.75 | Revise |
| 6 | Offal (liver or others) | 0.75 | 0.50 | Revise |
| 7 | Homemade sweets/cookies/candies/ice creams | 0.75 | 0.50 | Revise |
| 8 | Homemade sauces/condiments (chili, homemade sauces, dyes, bulk spices) | 0.25 | 0.25 | Reformulate |
| 9 | Street food (fried snacks, empanadas, sausages, juices sold in the street) | 0.50 | 0.00 | Reformulate |
| 10 | Canned foods (tuna, sardines, vegetables) | 0.75 | 0.25 | Revise |
| 11 | Packaged drinks with colorants (sodas, juices) | 0.75 | 0.50 | Revise |
| 12 | Ice of unknown origin (in juices, shaved ice, frozen treats) | 0.75 | 0.50 | Revise |
| 13 | Dairy products (milk, cheese, yogurt) | 1.00 | 0.50 | Adequate |
| 14 | Red meat, poultry, or eggs | 0.50 | 0.50 | Reformulate |
| 15 | Fruits rich in vitamin C (citrus, guava, blackberry) | 0.50 | 0.25 | Reformulate |
| 16 | Vegetables rich in calcium or iron (broccoli, chard, spinach) | 0.75 | 0.25 | Revise |
| 17 | Tea or coffee with meals | 0.50 | 0.25 | Reformulate |
| 18 | Meals on an empty stomach (e.g., breakfast only with tea/bread, no protein) | 0.25 | 0.25 | Reformulate |
| 19 | Usual source of drinking/cooking water | 1.00 | 0.50 | Adequate |
| 20 | Use of glazed clay or handmade ceramic utensils for cooking/serving acidic foods | 0.75 | 0.25 | Revise |
| 21 | Use of aluminum pots or recycled containers for cooking | 0.75 | 0.25 | Revise |
| 22 | Weekly frequency with which the child eats outside the home | 0.75 | 0.25 | Revise |
| 23 | Main places where the child eats outside the home (school, cafeteria, street, restaurant, etc.) | 0.75 | 0.25 | Revise |

Six items with an I-CVI below 0.78 were identified, primarily in the absorption modulators subsection and the consumption locations section. These items were: (15) fruits rich in vitamin C, (16) vegetables rich in calcium or iron, (17) tea or coffee with meals, (18) eating on an empty stomach, (22) frequency of consumption outside the home, and (23) places of consumption outside the home.

In all cases, the experts recommended adjustments to the wording or expansion of local examples, rather than item removal.

The qualitative suggestions were grouped into three categories: (a) clarifying food examples (e.g., including flours in cereals or additional examples of organ meats) and (b) contextualizing cultural habits (e.g., expanding items on tea, coffee, or eating on an empty stomach). These observations were incorporated into the final version of Block B, preserving the original conceptual structure as can be seen in Table 3.

**Table 3.** Expert feedback and final adjustments for each item of Block B (v1.2): dietary pattern and potential lead exposure questionnaire.

| No. | Original item | Experts' comments / observations | Final adjusted version (v1.2) |
| --- | --- | --- | --- |
| 1 | Leafy vegetables (lettuce, spinach, parsley) | Considered clear and relevant. | No changes. |
| 2 | Root or tuber vegetables (potato, cassava, carrot, beet) | Adequate; maintain examples. | No changes. |
| 3 | Grains and cereals (rice, maize, bread/wheat, oats) | Suggested including "flours" for broader coverage. | Grains, cereals and flours (rice, maize, bread, oats, wheat or maize flour). |
| 4 | Legumes (beans, lentils, chickpeas, peas) | High consumption but low exposure; maintain as control item. | Legumes (beans, lentils, chickpeas, peas) no wording changes. |
| 5 | Fish, tuna, sardines, or seafood | Approved without observations. | No changes. |
| 6 | Offal (liver or others) | Add more examples of offal. | Offal (liver, kidney, gizzards or others). |
| 7 | Homemade sweets/cookies/candies/ice creams | Clear and relevant item. | No changes. |
| 8 | Homemade sauces/condiments (chili, homemade sauces, dyes, bulk spices) | Adequate; sufficient clarity. | No changes. |
| 9 | Street food (fried snacks, empanadas, sausages, street juices) | Highly relevant; maintain examples. | No changes. |
| 10 | Canned foods (tuna, sardines, vegetables) | Add "fruits in syrup" as an additional example. | Canned or preserved foods (tuna, sardines, vegetables, fruits). |
| 11 | Packaged drinks with colorants (sodas, juices) | Clear and relevant item. | No changes. |
| 12 | Ice of unknown origin (in juices, shaved ice, frozen treats) | Maintain; important source of exposure. | No changes. |
| 13 | Dairy products (milk, cheese, yogurt) | No observations; relevant as modulator. | No changes. |
| 14 | Red meat, poultry or eggs | Relevant; no observations. | No changes. |
| 15 | Fruits rich in vitamin C (citrus, guava, blackberry) | Expand examples and add “other common fruits.” | Fruits rich in vitamin C (orange, lemon, guava, blackberry or other fresh fruits). |
| 16 | Vegetables rich in calcium or iron (broccoli, chard, spinach) | Avoid repetition with item 1; broaden examples. | Vegetables rich in calcium or iron (broccoli, kale, spinach, asparagus or beans). |
| 17 | Tea or coffee with meals | Add “during the day.” | Tea or coffee with meals or during the day. |
| 18 | Meals on an empty stomach (e.g., breakfast only with tea/bread, no protein) | Include other meals (lunch or dinner). | Meals on an empty stomach (e.g., breakfast, lunch or dinner only with tea/bread, without protein such as meat or eggs). |
| 19 | Usual source of drinking/cooking water | Add detail on whether the water is filtered or stored. | Usual source of drinking or cooking water (indicate if filtered, bottled, piped, or well water). |
| 20 | Use of glazed clay or handmade ceramic utensils for cooking/serving acidic foods | No relevant changes. | No changes. |
| 21 | Use of aluminum pots or recycled containers for cooking | Add “food thermos or lunch containers.” | Use of aluminum pots, recycled containers, food thermos or lunch containers. |
| 22 | Weekly frequency with which the child eats outside the home | Confusion about time frame (weekly vs monthly); unify with 30-day frame. | During the last 30 days, how often did the child eat outside the home (e.g., in the street, at school, or in a cafeteria)? |
| 23 | Main places where the child eats outside the home (school, cafeteria, street, restaurant, etc.) | Add more response options. | Main places where the child eats outside the home (select all applicable: school, cafeteria, street, restaurant or others). |

The questions added to the questionnaire, which will be sent via Google Forms to the representatives of the participating children (S1), were designed to obtain descriptive information about the children’s food consumption, rather than to create a construct or scale. Their validation focused exclusively on the clarity and comprehensibility of each question. Therefore, it was not necessary to evaluate psychometric properties such as internal consistency or construct validity.

### Statistical analysis

The statistical analysis will focus on (WISC-V) as the primary outcome and lead concentration in enamel as the main exposure, modeling using linear regression, with adjustments for age, sex, and other social and demographic variables collected in the consumption pattern questionnaire (e.g., maternal education/income), and fixed school effects with cluster-robust standard errors by school.

As secondary analyses, models will be repeated for WISC-V indices (VCI, VSI, FRI, WMI, PSI) and IQ and lead means will be compared between schools/sectors using ANOVA or Kruskal–Wallis,

For description, means and standard deviation or medians and interquartile ranges and proportions will be reported, and the main results as β with 95% CI and p. The statistical program SPSS version 25 will be used.

## Ethical statement

This project was reviewed and approved by the Human Research Ethics Committee of the Universidad de Especialidades Espíritu Santo under code C-UEES-25-06. This committee is authorized by the Ministry of Public Health to approve research protocols in Ecuador with the code DNIVS-CEISH-09-UEES-44 and is also registered with the Office for Human Research Protections of the United States under number IRB00014797.

The data collection process has not yet begun. Currently, approval has been obtained from the participating institutions, and data collection plans have been developed. Specifically, the most suitable locations for data collection in each school have been identified, along with the estimated timeframe for the process. Informed consent forms will be sent digitally to the children’s legal guardians via a link in the Google Form containing the consumption pattern survey. Only children with parental/guardian authorization to participate will be considered. Data collection is expected to begin in March 2026 and conclude in April 2026.

To ensure that participants understand the purpose, procedures, and potential risks of the study, the entire protocol will be explained and proof of understanding will be requested through the signing of the informed consent form. All participants will have the option to refuse to participate in any phase of the research without penalty. Participants will not receive any financial or academic compensation, nor will they pay anything for the analyses that will be performed in this research.

## Justification

Children’s exposure to lead remains one of the most important and preventable threats to neurodevelopment worldwide. According to the World Health Organization (WHO), even low-level exposure to lead can have irreversible effects on children’s developing brains, associated with lower intelligence quotient (IQ), attention-span deficits and diminished educational attainment [7]. Meta-analyses of data on lead in children’s blood have consistently demonstrated an inverse dose-response relationship between blood lead concentration and IQ, with IQ drops of several points for each increase in exposure, highlighting that a safe threshold has not yet been clearly defined. [37]. These findings underscore the urgent need for further investigation of lead exposure pathways and their cognitive consequences, especially in settings where regulatory and exposure controls may be weaker and where dietary and environmental sources of lead may differ from high-income countries.

In Ecuador, and particularly in urban and periurban areas, the true burden of childhood lead exposure remains largely undocumented. Few studies have measured lead biomarkers in children, and those that exist point to worrying levels of contamination. For instance, research in Andean mining zones of Ecuador found that infants and young children had some of the highest blood lead concentrations reported globally, linked to long-term environmental exposure from ore-processing activities [19]. Previous studies have also shown significant contamination of vegetables and proteins sold in easily accessible markets in large cities such as Quito, Ecuador [18]. However, no studies have systematically evaluated lead exposure among children in densely populated urban settings such as Guayaquil, nor have they investigated dietary and domestic factors, such as artisanal cookware, street food, or unsafe water sources, that may contribute to chronic, low-level exposure. This lack of surveillance and integrated exposure assessment underscores the urgent need for epidemiological studies that combine objective biomarkers of lead with dietary and environmental data to inform evidence-based prevention strategies in Ecuador.

Despite the established link between lead exposure and cognitive impairment in children from high-income countries, significant gaps remain in the evidence base for low- and middle-income settings. Recent meta-analyses indicate that even very low blood lead levels are associated with reduced IQ and learning outcomes [38] and that mean levels of exposure in 34 low- and middle-income countries vary significantly (e.g., 1.7–9.3 μg/dL) with potentially larger cognitive effects [39]. However, few studies have employed biomarkers of cumulative lead exposure (such as dental enamel) nor combined these with detailed dietary and household exposure data; even fewer have been conducted in Latin American urban contexts such as Ecuador. This paucity of data limits our ability to derive locally-relevant dose–response relationships or to identify modifiable exposure pathways that could be targeted for intervention.

This study will provide a novel contribution to environmental health research in Latin America by integrating cognitive assessment, dietary evaluation, and enamel biomarker analysis within a single framework. Recent evidence has shown that dietary exposure represents an important but often under-recognized pathway for lead intake in children, with studies identifying food as a dominant contributor to total exposure, particularly in low- and middle-income countries [30,40]. The use of tooth-based biomarkers such as enamel or dentine provides a reliable indicator of cumulative lead exposure across early developmental periods and correlates strongly with long-term neurocognitive outcomes [41]. By combining enamel lead quantification with a validated dietary instrument (DQQ-Ecuador and the Food Exposure Module), the study will quantify cumulative lead exposure and describe dietary factors potentially relevant to lead exposure pathways, including calcium and vitamin C intake, and assess their associations with enamel lead concentration. Using school-cluster modeling and multivariable regression, the analyses will estimate the association between cumulative lead exposure and IQ, contributing evidence from and under-studied geographical settings and addressing a key gap in the literature on child neurodevelopment and environmental toxicants.

The study aims to provide locally relevant data on cumulative lead exposure and describe its relationship with neurocognitive outcomes in Ecuador. The results may contribute to the evidence base used in policy and educational planning in similar middle-income settings. Recent analyses have emphasized that even modest reductions in childhood lead exposure yield measurable societal benefits, including higher lifetime earnings and improved educational outcomes [42]. Evidence from multi-country modeling studies indicates that the global economic cost of childhood lead exposure exceeds one trillion dollars annually, disproportionately affecting low- and middle-income countries [43]. Understanding the local contribution of dietary and domestic sources to lead burden will therefore enable targeted prevention measures, such as monitoring of food contact materials, safer artisanal production, and nutritional interventions aimed at reducing lead absorption.

Similar protocols linking biomonitoring and cognitive assessment have demonstrated that community-based, school-centered data collection can yield robust and generalizable results in resource limited settings [44,45]. Building on these precedents, the present study advances a contextually adapted, multidisciplinary design that can serve as a reference for future research on heavy metals and child neurodevelopment in Latin America.

## Limitations

Among the main limitations that can be anticipated in this research protocol, we can mention that the children’s IQ cannot be causally related to the level of lead found in their tooth enamel. There are other variables that could explain this level.

The questionnaire used to correlates food consumption patterns is subject to memory bias and possible biased responses from parents for various reasons.

## Dissemination plans

The protocol and research results will be disseminated through publications in scientific journals. In the event of modifications during the protocol review process, these will be adapted to the questionnaire and data collection procedure.

## Data Availability

No datasets were generated or analysed during the current study. All relevant data from this study will be made available upon study completion.

## Supporting information

Supplementary File 1. Google Form with the modifications made in the content validation process by experts.

Supplementary File 2. Expert assessment regarding the relevance, clarity and decisiveness of the questions added to the form.

## Acknowledgments

We would like to thank the Research Center of the Universidad Espíritu Santo for their trust and support in the creation of this research protocol. We also thank the Guayaquil Archdiocese Educational Network and all the schools participating in the project, especially their administrators and psychologists who helped us coordinate the necessary actions for this project to be carried out.

## Autor contributions

**Conceptualization:** Marcelo Armijos Briones; Diana Lucía Tinoco Caicedo; Nicole Armijos Bazurto; Brenda Angélica Bucheli

**Methodology**: Marcelo Armijos Briones; Diana Lucía Tinoco Caicedo; Nicole Armijos Bazurto; Brenda Angélica Bucheli; Elisa Iturralde Brinkmann; Dana Milena Crespo Párraga; Patricia Andrea Uscocovich Jiménez.

**Project Administration:** Marcelo Armijos Briones; Nicole Armijos Bazurto

**Validation:** Diana Lucía Tinoco-Caicedo; Paola Calle-Delgado; Christian Moreno Alvarado^2^; Diana Glicelia Tomalá Castro^4^; Paula Viktoria Báez Freire; Emily Angeline Zambrano Mendoza; Doménica Barcia Roca

**Writing – Original Draft Preparation:** Marcelo Armijos Briones; Elisa Iturralde Brinkmann; Dana Milena Crespo Párraga; Diana Glicelia Tomalá Castro; Paula Viktoria Báez Freire; Emily Angeline Zambrano Mendoza; Doménica Barcia Roca

